# Comparing Deep Learning Models for Lung Nodule Detection on Photon-Counting CT Images in a Non-Screening Setting

**DOI:** 10.64898/2026.09.28.26364159

**Authors:** Leonie Thieme, Eike Petersen, Kai Geissler, Laurens Müller-Groh, Steffen Oeltze-Jafra, Hoen-oh Shin, Sarah C. Scharm, Frank Wacker, Zahra Ahmadi, Andrea Schenk

## Abstract

**Background:** Comprehensive evaluations of lung nodule detection methods remain limited, particularly regarding data from clinical routine, novel photon-counting CT (PCCT) technology, and the comparison of academic and commercial models.

**Materials and Methods:** This retrospective study collected 1860 routine PCCT scans acquired at Hannover Medical School from 2021 to 2024. We compared four deep-learning-based lung nodule detection models, which were developed for conventional CT, namely the publicly available TotalSegmentator, nnDetection, and RadYOLO methods, as well as a commercial lung computer-aided detection (CAD) system. We evaluated the inter-model agreement in an unannotated cohort (n=1699), sensitivity in a smaller cohort of 25 manually annotated scans, and false positives as detections in 136 report-negative scans.

**Results:** In the unannotated cohort, the commercial CAD system detected 7071, TotalSegmentator 4264, nnDetection 5254, and RadYOLO 3122 nodules. Overall, the inter-model agreement was low, and the models produced substantially different candidate nodules. Approximately 1300 nodules were detected by all models, representing 18.7% - 42.8% of each model’s detections. 15.4% - 37.4% of a model’s detections did not match any other model. The models showed moderate sensitivity (between 0.71, 95%-CI [0.64, 0.77], and 0.83, 95%-CI [0.77, 0.88]), and TotalSegmentator and RadYOLO produced few false positives per scan (both 0.85, 95%-CI [0.70, 1.02]).

**Conclusion:** The lung nodules detected by four deep learning models showed high variation and a lack of consensus on clinical PCCT data. These findings highlight the importance of model benchmarking and local validation when deploying models in real-world clinical workflows.

## 1. Introduction

Lung cancer is the leading cause of cancerrelated mortality worldwide with an estimated 1.8 million deaths in 2022 [1]. It is often asymptomatic in early stages, however, early detection is crucial for successful treatment. A common sign of early-stage lung cancer are lung nodules, which are small, potentially malignant lesions in the lung [2]. In clinical practice, lung nodules are often discovered incidentally in computed tomography (CT) scans [2].

Artificial intelligence (AI)-based systems can support radiologists in nodule detection and management and are already being used in clinical practice [3]. Several deep learning models have been proposed and evaluated in low-dose screening or challenge settings [4, 5, 6, 7]. A recent study found 14 commercially available AI systems for lung nodule detection in screening programs with regulatory (CE) clearance in Europe [8], highlighting the importance of systematic benchmarking of different systems.

However, there is a lack of evaluation regarding incidental lung nodules in clinical routine data [9], where the variability in population characteristics and imaging parameters is higher than in standardized lung cancer screening programs. Evaluating the safety and effectiveness of radiological AI systems in the target setting is crucial [9, 10].

Existing validation studies on routine clinical data usually focus on either a single model or only commercially available systems [11, 12, 13, 14], limiting a comprehensive comparison across diverse deep learning approaches. In particular, to the best of our knowledge, no recent study has compared a currently available commercial system with common academic benchmark models.

In addition, AI-based lung nodule detection approaches have not been evaluated on novel photon-counting CT (PCCT) data outside phantom studies so far [15]. Unlike conventional energy-integrating CT detectors, PCCT directly converts X-ray photons into an electrical signal [16]. This results in a high dose efficiency and improved image quality due to increased spatial resolution and image contrast compared to conventional CT [16]. The PCCT technology has the potential to significantly benefit the clinical use of CT, including lung cancer diagnosis [16, 17, 18]. On phantom test data, an AI system trained on energy-integrating CT images achieved comparable sensitivity for nodule detection on PCCT and dose-matched energy-integrating CT [19]. This motivates the model transfer, however, whether detection models developed on conventional CT generalize to PCCT in clinical practice remains unclear.

This study performed a comparative analysis of three publicly available models and one commercial system for lung nodule detection using PCCT data collected during clinical routine. We quantified the overall inter-model agreement across a large unannotated clinical cohort by comparing the overlap between the detected nodules. Additionally, sensitivity and detections per report-negative scan were estimated on two smaller datasets.

## 2. Materials and Methods

The study was approved by the ethics committee at Hannover Medical School (No. 11510_BO_K_2024).

### 2.1. Data

We retrospectively collected routine chest and chest-abdomen PCCT scans of adult patients (≥18 years old) at Hannover Medical School, Germany. Images were acquired from 10/2021 to 11/2024 on a clinical PCCT system (NAEOTOM Alpha, Siemens Healthineers, Forchheim, Germany) with a reconstructed slice thickness of 1 mm. All scans are reconstructed using a soft Br40 kernel to reduce noise. In subset C (see section 2.1.3), we additionally use corresponding images that were reconstructed with a Br60 lung kernel. Appendix A.1 provides additional imaging parameters.

Since the manual annotation of all collected PCCT scans was not feasible, we used a large unannotated dataset (A) to quantify the inter-model agreement. For further experiments, we collected two smaller, clinically defined datasets with annotated nodules (B) and without clinically reported nodules (C) to assess sensitivity and detections in report-negative scans, respectively. In total, the study included 1860 scans of 1393 patients. An overview of the cohort characteristics is presented in Table 1.

**Table 1:** Cohort characteristics. **Dataset A**: unannotated agreement cohort (Sec. 2.1.1). **Dataset B**: cohort with manually annotated nodules (Sec. 2.1.2). **Dataset C**: cohort without clinically reported nodules and without cancer diagnoses (Sec. 2.1.3).

|  | A | B | C |
| --- | --- | --- | --- |
| Reported nodules | Unknown | Manual annotation | Report-negative |
| # Nodules | - | 217 | 0 reported |
| solid | - | 185 | - |
| report-based | - | 44 | - |
| sub-solid | - | 32 | - |
| # Scans | 1699 | 25 | 136 |
| # Patients | 1254 | 18 | 130 |
| cancer | 933 / 74% | 13 / 72% | 0 |
| lung cancer | 260 / 21% | 10 / 56% | 0 |
| primary | 154 / 12% | 8 / 44% | 0 |
| secondary | 112 / 9% | 2 / 11% | 0 |
| Age | 64.15 ± 13.93 | 63.80 ± 15.78 | 58.02 ± 17.55 |
| Sex | 540 / 43% F | 11 / 61% F | 47 / 36% F |
|  | 714 / 57% M | 7 / 39% M | 83 / 64% M |

#### 2.1.1. Unannotated data without nodule status information (Dataset A)

Dataset A included all 1699 eligible PCCT scans that were not assigned to datasets B or C. This dataset was unannotated and did not contain information about the nodule status.

#### 2.1.2. Data with nodule annotations (Dataset B)

Dataset B included PCCT scans of patients who underwent pathological examination of lung tissue (n = 25). Radiology reports were available from clinical routine. The scans were reviewed by a resident in radiology with 5 years of experience (S. C. S.), who manually detected and segmented lung nodules using watershed segmentation [20]. Only nodules with a diameter of 3 – 30 mm were included. In addition, we identified a subset of clinically relevant solid nodules that were explicitly mentioned in the radiology reports. A complete lesion annotation and reliable delineation could not be achieved in scans with disseminated nodules or lesions inseparable from tumour, inflammatory, or consolidative changes. Therefore, dataset B did not provide a complete reference for false-positive estimation.

#### 2.1.3. Data without nodules (Dataset C)

Dataset C included PCCT scans whose reports state the absence of clinically relevant nodules (n = 136; details in Appendix A.2). Patients with other nodular findings or a cancer diagnosis were excluded from this subset.

### 2.2. Nodule detection models

We compared four representative deeplearning-based models for lung nodule detection. We applied the academic and openly available TotalSegmentator, nnDetection, and RadYOLO model retrospectively. For the commercial computer-aided diagnosis (CAD) system (syngo.CT Lung CAD, Siemens Healthineers, Forchheim, Germany), which is used in clinical routine at Hannover Medical School, we extracted the results from the clinical systems.

#### 2.2.1. TotalSegmentator

TotalSegmentator is a publicly available set of pretrained models for the segmentation of anatomic structures in CT images [21]. The architecture is based on the state-of-the-art medical segmentation method nnU-Net, which automatically adapts a U-Net pipeline to a given dataset [22, 23]. Recently, a lung nodule segmentation model was added [21, 24], which was trained on 1353 subjects that are partly from the LIDC-IDRI dataset [25]. The model produces voxel-wise segmentation masks (Fig. 1a).

**Figure 1:**
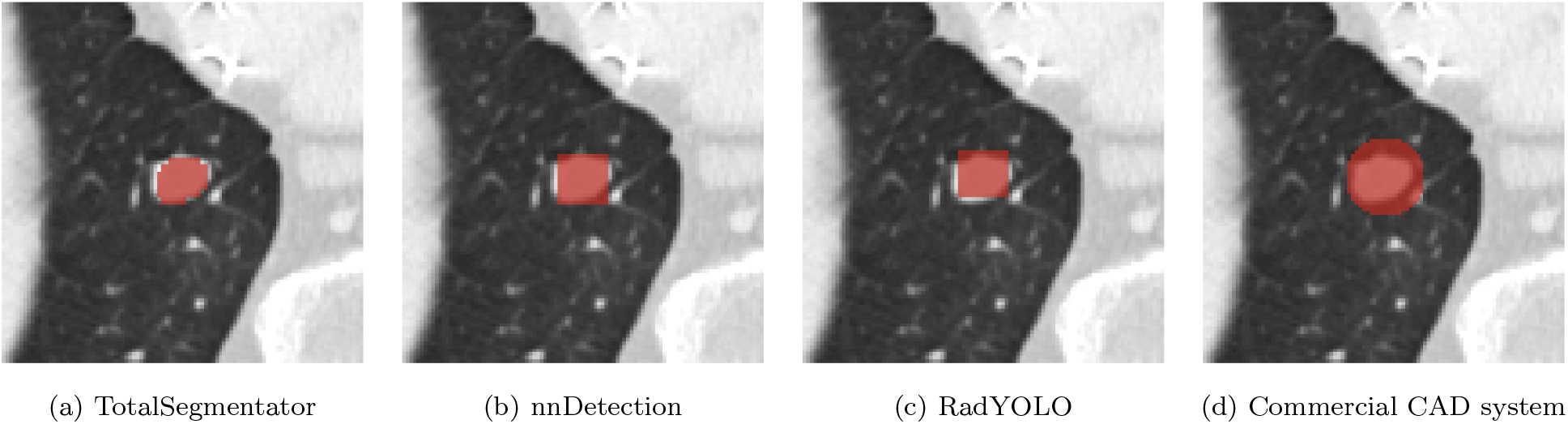
Example photon-counting CT image with overlaid nodule masks (red). (a) TotalSegmentator generates a voxel-wise segmentation mask and (b) nnDetection and (c) RadYOLO provide the coordinates of a bounding box, which are transformed into masks for visualization purposes only. (d) We extract a spherical mask from the original CAD image, which provides a circle without information on the nodule size.

#### 2.2.2. nnDetection

The publicly available nnDetection method follows nnU-Net’s self-configuring approach for medical images, but adapts the architecture to object detection [26]. Its effectiveness in lung nodule detection has been demonstrated on the popular LUNA16 benchmark [26, 27], a subset of the LIDC-IDRI dataset containing 888 CT scans. We trained the nnDetection model on LUNA16 using the original 10-fold cross-validation procedure^1^ and adapted the default inference parameters (Appendix B.1). The model produces 3D bounding box coordinates (Fig. 1b).

#### 2.2.3. RadYOLO

The publicly available RadYOLO model is based on the You Only Look Once (YOLO) architecture version 11 [28], an efficient object detection model for 2D images. RadYOLO extends the original YOLO model to 3D images by replacing all 2D image operations with their 3D analogs [29]. Appendix B.2 contains further model details. Like the nnDetection model, we trained RadYOLO from scratch on the LUNA16 dataset. The model produces 3D bounding box coordinates (Fig. 1c). We evaluated confidence thresholds of 0.1, 0.01, and 0.005 to balance between true and false positive rate. If not specified, RadYOLO refers to a confidence of 0.01, which yielded approximately one false positive per scan. This is seen as clinically acceptable [11].

#### 2.2.4. Commercial CAD system

The commercial system used was syngo.CT Lung CAD (versions VD20C/VD20D/ VD30A, Siemens Healthineers, Forchheim, Germany), which is integrated into the clinical workflow at Hannover Medical School for the detection of solid and sub-solid lung nodules. The system also provides a “solid-only” mode, which excludes suspected sub-solid and fully calcified findings.

Rather than lesion coordinates, the CAD system outputs graphical markers, whose size does not represent the lesion size. We extracted the coordinates of spherical nodule marker masks using thresholding and Hough-circle detection (Fig. 1d, Appendix B.3). Extraction was manually verified in the 411 CAD images corresponding to scans in dataset B, and no errors were observed.

### 2.3. Evaluation

We excluded detections outside the lung using lung segmentation masks generated by the total-body segmentation model of TotalSegmentator [21, 22]. We applied morphological closing and dilation to slightly expand the segmentation and close holes. Nodule detections were converted to bounding boxes and expanded to a minimum extent of 5 mm to reduce matching uncertainty for small lesions. Detection centers are defined as the midpoints of axis-aligned bounding boxes.

#### 2.3.1. Experiment A: inter-model agreement

The primary experiment A was designed as an inter-model agreement analysis on a large cohort (dataset A). The result of a model comparison was considered a directional match when the center of a detection from model 1 lies inside a detection from model 2 (Fig. 2a). We manually verified this procedure for 100 detected nodules.

**Figure 2:**
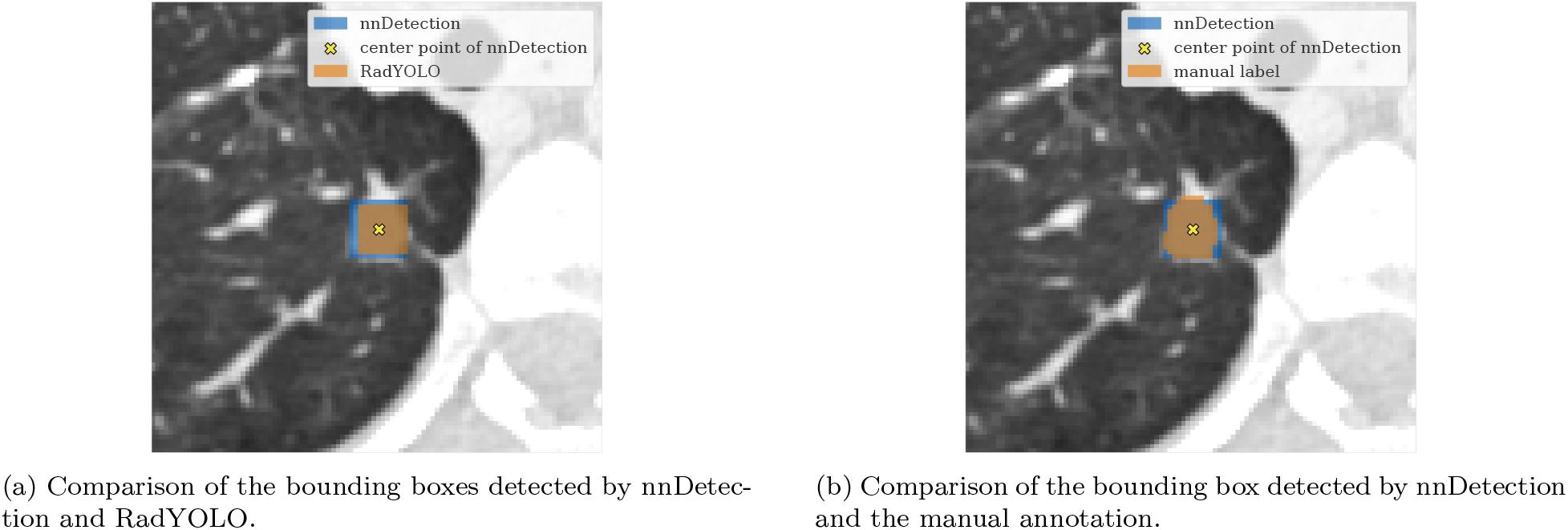
Center-point matching procedure. (a) The nnDetection candidate nodule is matched to RadYOLO because its center lies within the RadYOLO bounding box. (b) The nnDetection candidate nodule is counted as a true positive because its center lies within the manually labeled segmentation mask. Boxes are converted to masks only for visualization.

### 2.3.2. Experiment B: sensitivity

Experiment B evaluated the sensitivity for all manually annotated solid lung nodules and the clinically reported subset in dataset B. This analysis was limited to solid nodules due to the commercial CAD system’s “solid-only” mode. We calculated overall nodule-level sensitivity and mean sensitivity per scan, which is more robust to differences in the number of nodules per scan. A detected nodule was counted as a true positive if its center point lies within the reference segmentation mask (Fig. 2b). The 95%-confidence interval (95%-CI) of the sensitivity was calculated using the Agresti-Coull interval [30]. Because annotations were not exhaustive, additional detections by the AI models were not classified as false positives.

#### 2.3.3. Experiment C: detections per report-negative scan

Experiment C estimated the false positives per report-negative scan (FP/RNS) as the detections on dataset C, which includes images with no clinically reported nodules. The 95%-CI for a rate was calculated using the exact interval [31]. In addition, the impact of the reconstruction kernel choice was evaluated by comparing FP/RNS on Br40 and Br60 data.

## 3. Results

A total of 1393 patients and 1860 PCCT scans were included in this study. The dataset characteristics are displayed in Table 1.

### 3.1. Experiment A: inter-model agreement

The results of the inter-model agreement on dataset A are shown in Table 2. The pairwise model matching depends on the direction of the comparison, because size differences between overlapping boxes can result in the center point of detection 1 lying inside the detection 2, but not the other way around. Overall, the agreement between the different models was low. Of all detected nodules, only 1319 – 1353 detections in 1699 images were shared by all four models, which corresponds to 18.7% – 42.8% of each model’s detections. Model-unique detections occurred frequently for all models and ranged from 15.4% (808 nodules) for nnDetection to 37.4% (2646 nodules) for the commercial CAD system. Overall, the commercial CAD system detects the most nodules (7071) and RadYOLO the least (3122), which influences the high number of matches with the commercial CAD system and the low number of matches with RadYOLO for other models.

**Table 2:**
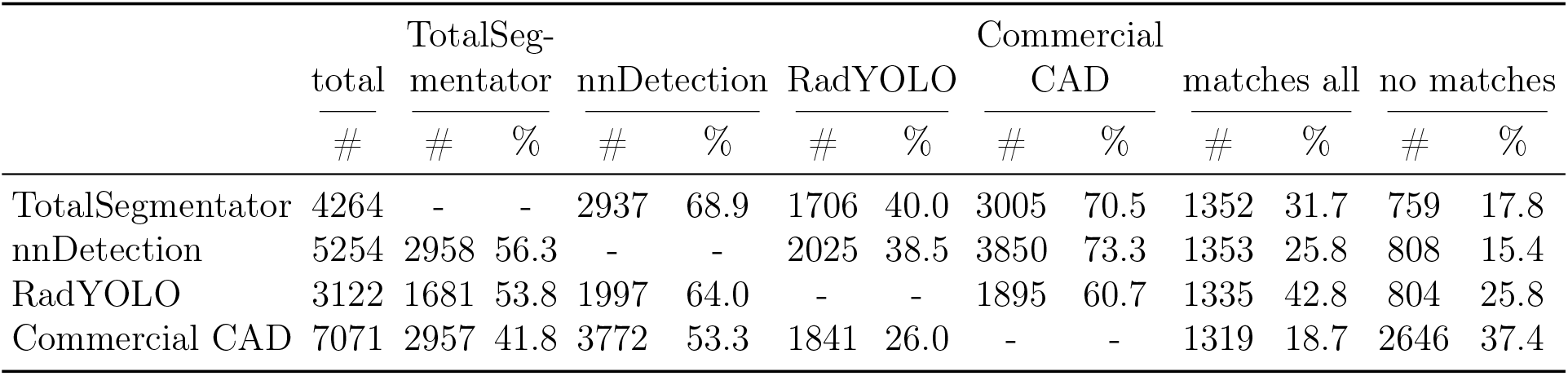
Total number of detected nodules per model and results of the directional inter-model agreement on the 1699 scans of dataset A. Percentages use each the respective model’s total detections as the denominator. The columns “matches all” and “no matches” represent detections that are shared with all or none of the other models, respectively.

Fig. 3 shows the number of detections that match at least one or two other models. Despite a lower number of total detections, the RadYOLO model showed a lower ratio of matches with at least one model (74.2%) than nnDetection (84.6%) and TotalSegmentator (82.2%). The largest three-model overlap was observed between TotalSegmentator, nnDetection, and the commercial CAD system, with a mean of 1154 matches compared to 69 – 298 for the other combinations.

**Figure 3:**
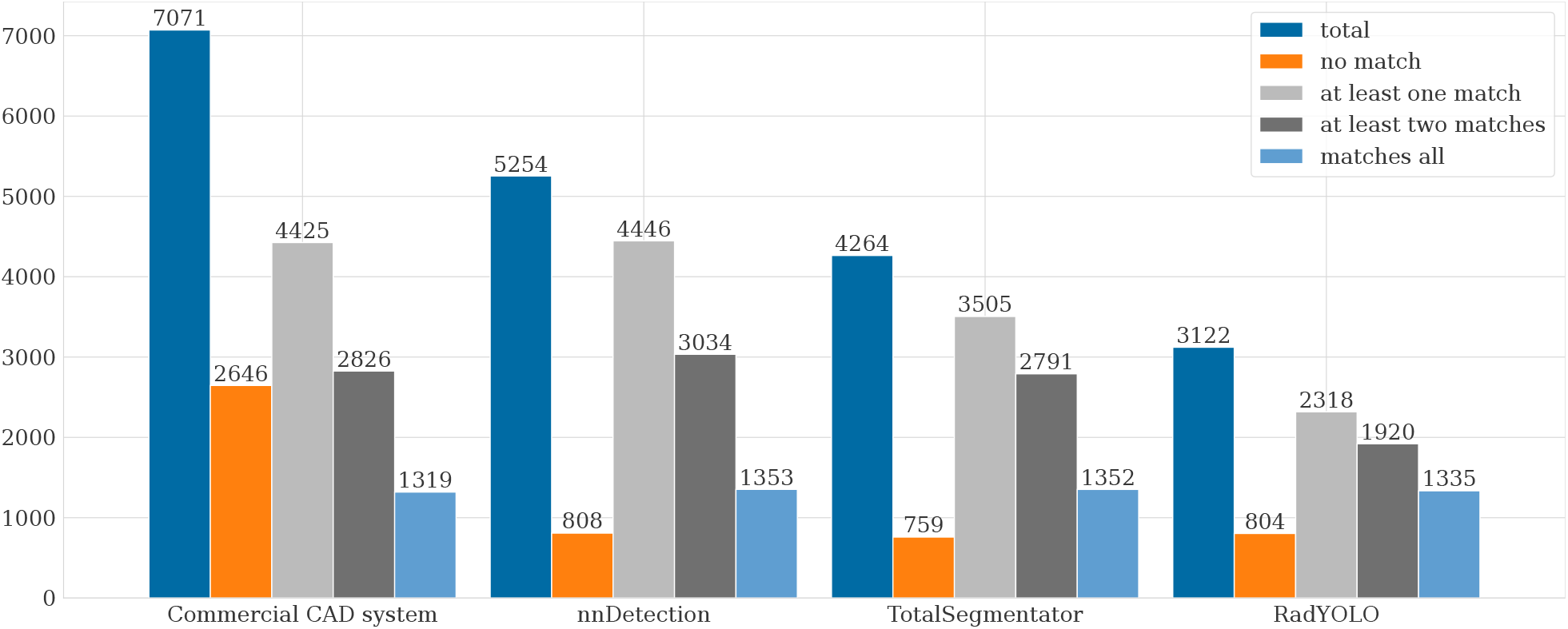
Inter-model agreement in dataset A. Bars show (from left to right) total detections, unmatched detections, and detections matched by at least one, at least two, or all three other models.

The sizes of the detected nodules differed across models, with median maximum extents of 4.1 mm for TotalSegmentator, 5.6 mm for nnDetection, and 7.7 mm for RadYOLO (Fig. 4). The commercial CAD system was excluded from this analysis because it does not provide nodule size information. For each model, nodule sizes were similar between all detected nodules (Fig. 4a) and nodules shared by all models (Fig. 4b), and no strong systematic shift was observed between the distributions. Therefore, there is no clear indication that a specific nodule size may be easier to detect by all models. It also shows that no model systematically detected more nodules of a specific size.

**Figure 4:**
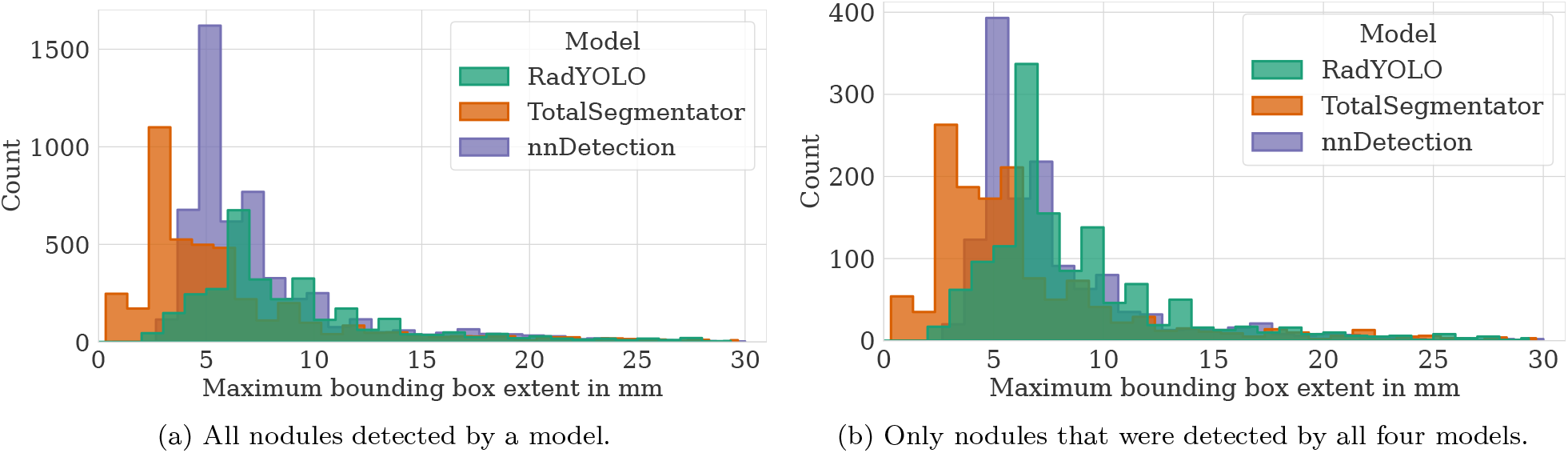
Maximum bounding box extent for (a) all detected nodules and (b) detected nodules shared by all four models in dataset A. The commercial CAD system is excluded in the chart because it provides no lesion-size information. The x-axes are truncated at 30 mm to enhance visibility.

### 3.2. Experiment B: sensitivity

For all 185 manually labeled solid nodules of dataset B, TotalSegmentator, RadYOLO, and the commercial CAD system achieved similar moderate sensitivity results of 0.71–0.75 with overlapping 95%-confidence intervals (Table 3). The nnDetection model showed a sensitivity of 0.83 (95%-CI [0.77, 0.88]). The mean number of detected nodules that did not match the manually labeled nodules were 7.39/scan for TotalSegmentator, 8.17/scan for RadYOLO, 9.30/scan for nnDetection, and 13.96/scan for the commercial CAD system. All models showed a higher sensitivity for the subset of 44 clinically reported solid nodules compared to all nodules. Also, the model sensitivities were substantially lower for small nodules, as well as for sub-solid nodules compared to solid nodules (Appendix D).

**Table 3:**
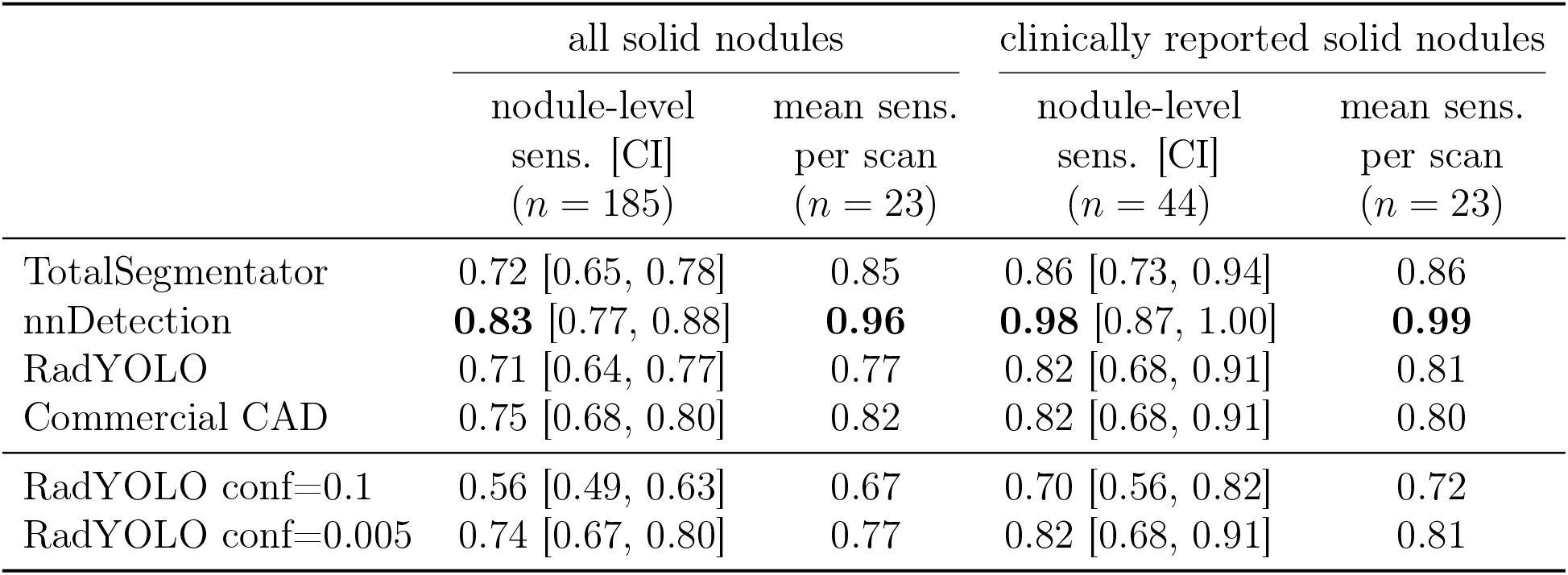
Nodule-level sensitivity (sens.) and mean sensitivity per scan for manually segmented solid nodules and the subset of clinically reported solid nodules in dataset B. The 95%-confidence intervals (CIs) are calculated using the Agresti-Coull interval [30]. The standard RadYOLO confidence parameter (conf) is 0.01.

### 3.3. Experiment C: false positives per report-negative scan

There was a trade-off between sensitivity and FP/RNS, which were both increased by lower RadYOLO confidence thresholds (Tables 3 and 4). A threshold of 0.01 matched approximately 1 false positive per scan, which is seen as clinically acceptable in the literature [11], and was used as the standard RadYOLO model. A similar trade-off between sensitivity and FP/RNS was observed during the testing of different inference parameters for the nnDetection model.

**Table 4:**
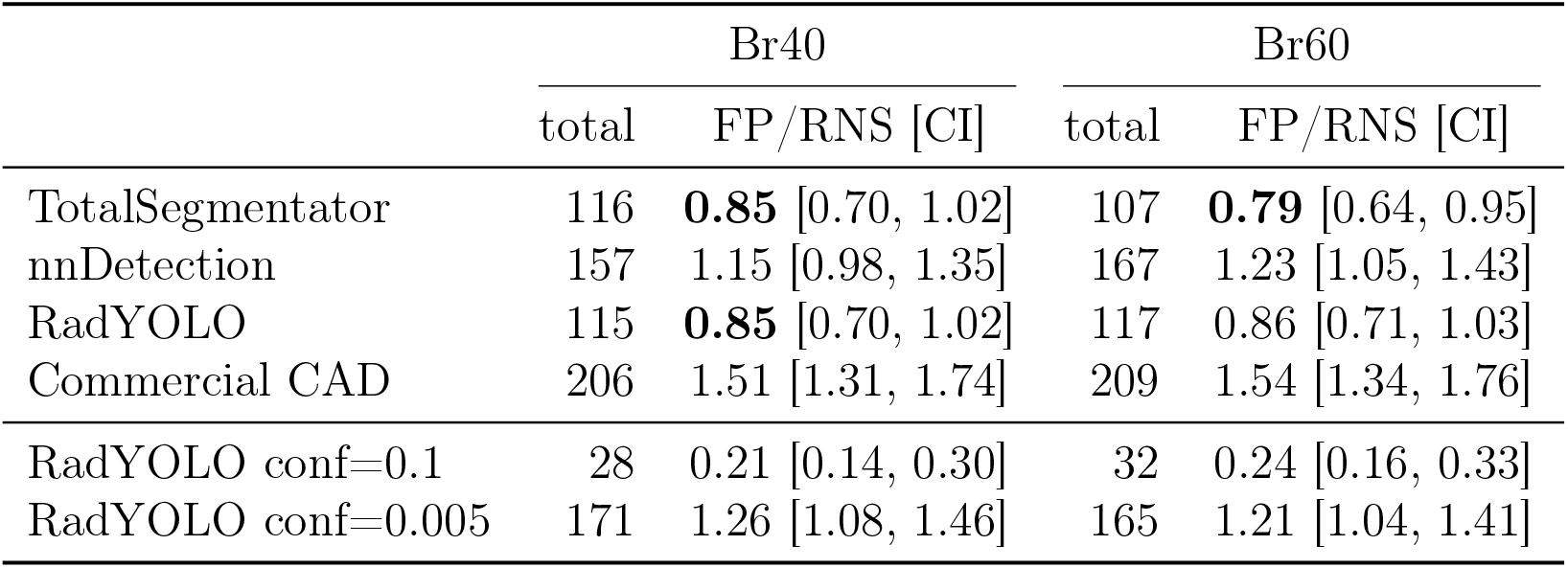
Total number of detected nodules and false positives per report-negative scan (FP/RNS) for 136 scans in dataset C reconstructed with Br40 or Br60 kernels. The 95%-confidence intervals (CIs) are calculated using the exact interval [31]. The standard RadYOLO confidence parameter (conf) is 0.01.

The TotalSegmentator and RadYOLO models achieved a low FP/RNS result (both 0.85 for Br40, see Table 4), which was below the threshold of 1 false positive per scan. The nnDetection model and the commercial CAD system showed a higher number of detections with 1.15 and 1.51 for Br40, respectively. We observed no substantial differences between the Br40 and Br60 reconstructions.

### 3.4. Inference Time

The RadYOLO model had the lowest mean inference time of 2.9 s per image, compared to 60.7 s per image for TotalSegmentator and more than 18 minutes per image for nnDetection (Appendix C).

## 4. Discussion

There is a lack of external, comparative evaluation of lung nodule detection models on real-world clinical data, particularly in the context of modern imaging techniques such as PCCT. We found that the four tested models produced strongly differing results.

As the three open academic models were largely trained on the same data, we expected a high degree of agreement between them. Interestingly, our results do not support this initial hypothesis. TotalSegmentator’s training set was partly from LIDC-IDRI, while nnDetection and RadYOLO were both trained on the LUNA16 subset derived from LIDC-IDRI. Still, the agreement between nnDetection and RadYOLO remained limited at 38.5% and 64.0%, respectively. We hypothesize that these large differences are caused, at least in part, by the substantial architectural and algorithmic differences between the three models, providing a counterpoint to the often-claimed supremacy of data over algorithms.

The observed sensitivities are comparable to previous evaluation studies of deep learning models on clinical data [27], which reported values in the range of 65 – 94% at 1 false positive per scan [6, 7, 11, 32]. In lung nodule detection, there is an inherent trade-off between sensitivity and falsepositive rate [9]. The total number of detected nodules is influenced by inference parameters, which specify, for example, the confidence threshold or the number of detections per scan. Here, we had to adapt the nnDetection inference parameters to reflect scans with a higher number of nodules in dataset B compared to the training data. Further adapting the inference parameters to increase the thresholds for detected nodules may lead to a lower FP/RNS, but also a lower sensitivity result. In our study, nnDetection had a sensitivity of 0.83 at 1.15 FP/RNS, which was lower than on LUNA16, where we reproduced the reported sensitivity of 0.95 at 1.0 false positive per scan [26]. A similar performance degradation from the LUNA16 benchmark to clinical practice has been reported previously [7, 32]. This suggests that model performance estimated on curated benchmark data does not directly transfer to routine clinical CT with different patient populations and acquisition parameters or technologies, such as PCCT, and that external model evaluations such as presented here remain crucial.

Still, neither model failed completely despite the substantial distribution shifts between training and evaluation datasets. The training sets were primarily based on the LIDC-IDRI dataset [25] and its LUNA16 subset [27], a widely-used benchmark for lung nodule detection. These datasets contain conventional CT data (not PCCT) from the US before 2011. Generalizability could be further improved by incorporating additional training data from clinical routine in Germany or images acquired with a PCCT scanner.

To the best of our knowledge, no recent study has compared the performance of current academic models with commercial ones, rendering it difficult for researchers and clinicians alike to make informed choices. In our comparative benchmarking of both academic models and a commercial system in an external real-world evaluation, none of the four systems clearly outperforms the others. We observed substantial differences in operating characteristics, indicating that no model can be considered uniformly superior. Prior comparisons of different commercial CAD systems on conventional CTs similarly reported substantial variation in sensitivity [14, 33], detection counts [13], and volume estimates [33]. Differences in measured lung nodule volume can impact the malignancy risk assessment and subsequent clinical management [33, 13]. Our study also observed different output sizes for the same nodules, however, the models in this study were trained for detection rather than accurate volume measurement.

While lung nodule detection research remains active [3, 11], the focus has shifted from detection to malignancy risk estimation, for example in the recent LUNA25 challenge [34]. However, Herber et al. [14] concluded that AI-based models are not yet ready for stand-alone clinical use for the task of nodule malignancy classification. Our findings extend this evidence to PCCT and show that substantial model variability is already present in the preceding detection step, before malignancy classification or guideline-based management.

While the results did not determine which model is more clinically reliable, they still have clinical significance. Nodule detection is one step in the clinical nodule management workflow, and different candidate nodules may impact radiologist review, volumetric assessment, risk stratification, and follow-up imaging. An unreliable AI system can lead to delayed cancer diagnoses due to false negatives, or an increased burden for patients and clinicians due to the need for follow-up monitoring of false positives. The low level of inter-model agreement highlights the importance of local validation, ongoing performance monitoring, and continued radiologist oversight after implementing a system in the clinic [10].

Several limitations should be acknowledged. Dataset B contains a comparatively small number of cases with a reference standard, which led to wide confidence intervals and limited interpretability of the sensitivity results. Also, constructing dataset C based only on radiology reports introduces uncertainty. Detected nodules reflect detections in report-negative scans rather than absolute false positives. Potential variability and ambiguity of label information extracted from reports may decrease in the future when structured reporting is more widely adopted [35]. Finally, our findings reflect the specific model versions and thresholds used in this study. As shown in Appendix B.1 for nnDetection, the model parameters can have a substantial influence on the model performance.

## 5. Conclusion

We collected 1860 PCCT scans from clinical routine in three datasets. We compared four deep learning models for lung nodule detection, namely TotalSegmentator, nnDetection, RadYOLO, and a commercial CAD system (Siemens syngo.CT Lung CAD). Overall, the models generated substantially different sets of candidate nodules and showed low inter-model agreement. Our findings highlight the importance of benchmarking, local validation in the target real-world setting, and ongoing monitoring when deploying models in clinical workflows. This is particularly important under domain shifts, such as when transferring models developed on conventional CT to modern PCCT. In the future, we aim to expand the annotated dataset and incorporate PCCT data into the training data to increase model adaptation and generalizability.

## Data Availability

The authors do not have permission to share data.

## Acknowledgments

This research is supported by the Ministry of Science and Culture of Lower Saxony through funds from the Volkswagen Foundation’s zukunft.niedersachsen program for the ‘CAIMed – Lower Saxony Center for Artificial Intelligence and Causal Methods in Medicine’ project (grant no. ZN4257).

The authors acknowledge the Hannover Medical School for providing Enterprise Clinical Research Data Warehouse (ECRDW) and MHH-HPC resources and technical support that have contributed to the research results reported within this paper.

## Declaration of generative AI and AI-assisted technologies in the manuscript preparation process

AI tools were used solely for grammar checks and language polishing. The authors conducted all scientific content, data analysis, and interpretation, and take full responsibility for the content of the published article.

## Footnotes

1 https://github.com/MIC-DKFZ/nnDetection

