## Supplemental Material for "Comparing Deep Learning Models for Lung Nodule Detection on Photon-Counting CT Images in a Non-Screening Setting"

### Appendix A. Data details

#### A.1. General data properties

The PCCT images have a reconstructed slice thickness of 1 mm and a matrix size of 512×512, 768×768, or 1024×1024 pixels, depending on the CT protocol used and radiation exposure in auto-mode. The field of view was adapted to the patient size and the voxel size ranges from 0.35 to 0.98 mm. Spiral pitch factor was 0.6 –2.4, kVp 120 – 140 kV, tube current 65 –1062 mA, and revolution time 0.25 s. The selected CT scans were acquired using three different protocols for contrast agent scans of the lung: the standard protocol for contrast agent scans, a staging protocol for thorax/abdomen scans, and the protocol for suspected lung embolism. An iodine contrast agent with a concentration of 400 mg/ml and a volume of 35–149.5 ml (typical volume of 80 ml) was applied.

#### A.2. Data without nodules

To create a dataset of CT images without lung nodules, we manually compile a list of phrases that are commonly used by radiologists in a radiology report to describe the absence of lung nodules. We include cases that contain phrases from Supplemental Table A.1 in the corresponding (German) radiology report. The comparison is performed using regular expressions to include uppercase and lowercase and singular and plural forms. We exclude cases that mention other nodular structures with the keyword "nodul".

### Appendix B. Model details

#### B.1. nnDetection

The following inference parameters were used for the nnDetection model:

– model_score_thresh: 0.7
– ensemble_score_thresh: 0.7
– model_topk: 5000
– model_detections_per_image: 500
– ensemble_topk: 5000
– remove_small_boxes: 1.0 (in pixel space)

We adapted the default inference parameters (see below) to allow for more possible detections per image and cope with a high number of nodules present in a few of the collected CTs. When using the default inference parameters from LUNA16, nnDetection showed an under-detection of nodules in scans with many true nodules (#nodules > 25). This resulted in a low nodule-level sensitivity of 0.39 (95%-CI [0.32, 0.46]) on all solid nodules in dataset B. The default LUNA16 parameters tested in the beginning were:

– model_score_thresh: 0.5
– ensemble_score_thresh: 0.5
– model_topk: 1000
– model_detections_per_image: 100
– ensemble_topk: 1000
– remove_small_boxes: 4.0

| German | English translation |
|---|---|
| kein pulmonaler rundherd | no pulmonal nodule |
| kein pulmonaler lungenrundherd | no pulmonal lung nodule |
| kein suspekter rundherd | no suspect nodule |
| kein suspekter lungenrundherd | no suspect lung nodule |
| kein suspekter pulmonaler rundherd | no suspect pulmonal lung nodule |
| kein malignomsuspekter pulmonaler rundherd | no suspected malignant pulmonal nodule |
| kein malignomsuspekter rundherd | no suspected malignant nodule |
| kein malignomsuspekter lungenrundherd | no suspected malignant lung nodule |
| kein nachweis von tumorsuspekten pulmonalen rundherd | no evidence of tumour-suspicious pulmonary nodule |
| kein nachweis von tumorverdächtigen rundherd | no evidence of tumour-suspicious pulmonary nodule |
| kein nachweis von tumorverdächtigen lungenrundherd | no evidence of tumour-suspicious lung nodule |
| keine pulmonalen rundherd | no pulmonal nodule |
| keine suspekten rundherd | no suspect nodule |
| keine suspekten lungenrundherd | no suspect lung nodule |
| keine suspekten pulmonalen rundherd | no suspect pulmonal nodule |
| keine malignomsuspekten pulmonalen rundherd | no suspected malignant pulmonary nodules |
| keine malignomsuspekten rundherd | no suspected malignant nodules |
| keine malignomsuspekten lungenrundherd | no suspected malignant lung nodules |

Table A.1: Manually compiled list of phrases that are commonly used by radiologists in a radiology report to describe the absence of lung nodules. Provided are the original German phrases and an approximate English translation. The comparison is performed using regular expressions to include uppercase and lowercase and singular and plural forms.

#### *B.2. RadYOLO*

RadYOLO extends the original You Only Look Once (YOLO) architecture version 11 [28] to 3D imaging volumes by replacing all 2D layers, like convolution, pooling, and upsampling layers, with their 3D analogs [29]. The YOLO model performs a simultaneous evaluation of the entire image in one inference pass by dividing the image into a grid with independent cell predictions. The grid cells act on the model levels that are downsampled by a factor of 8, 16, and 32 for each axis. For each grid cell, a confidence score and a 3D bounding box are predicted. RadYOLO uses the 's' model size configuration, resamples the input image to a fixed resolution of $352 \times 352 \times 192$, and normalizes the image intensities with z-score normalization, where the intensity mean and standard deviation inside the reference boxes of the LUNA16 dataset are used. In addition, clipping to the 0.5 and 99.5 percentiles of the LUNA16 image intensities is applied. In contrast to the nnDetection and TotalSegmentator (which are based on nnU-Net) backbones, the RadYOLO architecture employs downsampling layers earlier in the neural network, which makes it less computationally and memory intensive.

In the final layer, six bounding box coordinates are predicted, instead of four. The bounding box losses (CIoU [36] and DFL [37]) are naturally extended to 3D when switching from 2D to 3D boxes. The reference bounding box assignment during training is done using the TaskAlignedAssigner [38], like in YOLO11, and box confidences are optimized using a binary cross entropy loss. It applies standard data augmentations like axis flipping, rotation, scaling, and intensity augmentations, similar to nnU-Net and nnDetection.

#### *B.3. Commercial CAD system*

The syngo.CT Lung CAD system (Siemens Healthineers, Forchheim, Germany) is integrated into the clinical workflow at Hannover Medical School and applied automatically to new CT scans. To enable the comparison of all four models, we selected images that have the result of the syngo.CT Lung CAD system available in the clinical system, including results where it detects no nodules. The CAD system outputs 2D images that contain a CT image slice with the nodule detection results ingrained on the image pixels. If necessary, we apply resampling to match the resolution between the CT image and CAD result. We extract the slice number of a CAD image based on the coordinates in the Image Position (Patient) DICOM attribute.

To extract the correct position of the detected nodules, we resolve slight differences between the output and the corresponding CT image, for example, regarding zoom. We find the scaling and translation transformation between the images using the Enhanced Correlation Coefficient algorithm implemented in OpenCV (cv2.findTransformECC, OpenCV version 4.10.0).

We apply a threshold to the image to generate a binary mask of the overlaid CAD results. We use the Hough circle transform for circle detection to extract coordinates of detected nodules [39]. The circles used as markers are of a similar size, irrespective of the lesion size. Therefore, we set the initial minimum and maximum radius of detected circles in the Hough transform based on the image size. The minimum radius threshold is increased if the number of detected nodules from the initial circle detection exceeds the target number of total detections in the current CAD output image, which is overlaid in the image as text. We extract the number of detected nodules from the text using the pytesseract package (version 0.3.13), an open-source wrapper for Google's optical character recognition (OCR) Tesseract engine. The resulting circle-shaped mask around the detected nodule is expanded to a 3D sphere. We manually verified the nodules extracted in this way in 411 CAD images corresponding to scans in dataset B and observed no errors regarding the extracted detections.

### Appendix C. Inference time

We calculate the inference time of TotalSegmentator, nnDetection, and Rad-YOLO on 380 input files with an average file size of 261.0 MB (± 51.1 MB). We use the default LUNA16 inference parameters for nnDetection (Appendix B.1). The commercial CAD system is not analysed because it is run prospectively in clinical routine. Calculations are performed using an NVIDIA A100 80 GB graphics card. The total time from input to output is measured, including pre- and post-processing.

The YOLO model family is known to be very fast [28]. This also applies to the 3D extension, which achieves the lowest inference time of 2.9 seconds per image on average (Supplemental Table C.1). In contrast, the nnDetection model requires an average of 25.1 seconds for pre-processing and 18 minutes to process one image due to its computationally expensive post-processing. The output bounding box probabilities have to be post-processed to cluster overlapping bounding boxes or remove boxes with very low prediction scores, for example [26]. An nnU-Net, such as the one used in TotalSegmentator, does not require these time-consuming post-processing steps. The TotalSegmentator model has an average inference time of 60.7 seconds.

### Appendix D. Additional experiments regarding nodule size and type

We performed additional analyses based on the sizes of the manually segmented solid nodules of dataset B, which are between 3 and 30 mm. The size bins are based on clinical guidelines for incidental nodule management [40]. The dataset contains 56 nodules with a size $< 6$ mm, 36 nodules 6 - 8 mm, and 93 nodules $> 8$ mm. The size is calculated as the maximum extent of the axis-aligned bounding box. All four models showed a substantially lower sensitivity for small nodules (Supplemental Table D.1), particularly on nodules with a size $< 6$ mm. The commercial CAD system showed comparatively robust results on smaller nodules.

We also evaluate the sensitivity based on the manually labeled nodule type in dataset B, which contains 185 solid and 32 sub-solid nodules (Supplemental Table D.2). The commercial CAD system is excluded because it is

| | mean [s] | stdev [s] |
|---|---|---|
| TotalSegmentator | 60.7 | 6.0 |
| nnDetection pre-processing | 25.1 | 18.7 |
| nnDetection model inference | 1080.3 | 333.0 |
| RadYOLO | **2.9** | **1.5** |

Table C.1: Mean and standard deviation (stdev) of the inference time per image in seconds for TotalSegmentator, nnDetection and RadYOLO. Calculations are performed on 380 PCCT scans using an NVIDIA A100 80 GB graphics card. The commercial CAD system is excluded from this analysis because it is run prospectively in clinical routine. For nnDetection, pre-processing and model inference time are reported separately, because the pre-processing procedure generates and saves intermediate results that can be reused for subsequent inference.

partially run in a "solid-only" mode, which excludes suspected sub-solid or fully calcified nodules. The LIDC-IDRI dataset, on which the TotalSegmentator, nnDetection, and RadYOLO models were at least partially trained, contains both solid and sub-solid nodules. All three models demonstrated a substantially lower sensitivity on sub-solid nodules compared to solid nodules.

| | $< 6\,\mathrm{mm}$ | | $6-8\,\mathrm{mm}$ | | $> 8\,\mathrm{mm}$ | |
|---|---|---|---|---|---|---|
| | nodule-level sens. [CI] ($n = 56$) | mean sens. per scan ($n = 11$) | nodule-level sens. [CI] ($n = 36$) | mean sens. per scan ($n = 8$) | nodule-level sens. [CI] ($n = 93$) | mean sens. per scan ($n = 19$) |
| TotalSegmentator | 0.39 [0.28, 0.52] | 0.57 | 0.83 [0.68, 0.93] | 0.95 | 0.87 [0.79, 0.93] | 0.94 |
| nnDetection | 0.66 [0.53, 0.77] | 0.85 | 0.83 [0.68, 0.93] | 0.96 | 0.92 [0.85, 0.97] | 0.99 |
| RadYOLO | 0.41 [0.29, 0.54] | 0.45 | 0.81 [0.65, 0.91] | 0.83 | 0.85 [0.76, 0.91] | 0.91 |
| Commercial CAD | 0.64 [0.51, 0.76] | 0.82 | 0.86 [0.71, 0.94] | 0.97 | 0.76 [0.67, 0.84] | 0.84 |

Table D.1: Nodule-level sensitivity (sens.) and mean sensitivity per scan when comparing model results to subsets of the manually segmented solid nodules from dataset B based on nodule size. The 95%-confidence interval (CI) of the sensitivity is calculated using the Agresti-Coull interval [30].

| | sub-solid | |
|---|---|---|
| | nodule-level sens. [CI] ($n = 32$) | mean sens. per scan ($n = 11$) |
| TotalSegmentator | 0.50 [0.34, 0.66] | 0.61 |
| nnDetection | 0.59 [0.42, 0.75] | 0.71 |
| RadYOLO | 0.59 [0.42, 0.75] | 0.71 |

Table D.2: Nodule-level sensitivity (sens.) and mean sensitivity per scan when comparing model results to sub-solid manually segmented nodules from dataset B. The 95%-confidence interval (CI) of the sensitivity is calculated using the Agresti-Coull interval [30].
